# The Alaska Native/American Indian experience of hepatitis C treatment with sofosbuvir-based direct-acting antivirals

**DOI:** 10.1101/2021.09.03.21263089

**Authors:** Lisa Townshend-Bulson, Elena Roik, Youssef Barbour, Dana Bruden, Chriss Homan, Hannah Espera, Tim Stevenson, Annette Hewitt, Wileina Rhodes, Jim Gove, Julia Plotnik, Mary Snowball, John McGilvray, Brenna C. Simons, Brian McMahon

## Abstract

**Background:** Direct-acting antiviral (DAA) drugs have been effective in the treatment of chronic hepatitis C virus (HCV) infection. Limited data are available on safety, tolerability, and efficacy in American Indian or Alaska Native people. We aim to evaluate the treatment outcomes of sofosbuvir-based regimens for treatment of HCV in a real life setting in Alaska Native/American Indian (AN/AI) people.

**Methods:** AN/AI patients within the Alaska Tribal Health System with confirmed positive anti-HCV and HCV RNA, who were 18 years of age and older were included in the study. Pretreatment baseline patient characteristics, treatment efficacy based on sustained virologic response (SVR) 12 weeks after treatment completion, and adverse effects were assessed. The following treatments were given according to the American Association for the Study of Liver Diseases/Infectious Disease Society of America (AASLD/IDSA) HCV Guidance: ledipasvir/sofosbuvir, sofosbuvir plus weight-based ribavirin, and sofosbuvir/velpatasvir.

**Results:** We included 501 patients with a mean age of 54.3 (range 21.3 -78.3) in the study. Overall SVR was achieved in 95.2% of patients who received one of the three DAA regimens. For those with cirrhosis, overall SVR was 92.8% and for those with genotype 3 91.1% achieved SVR. The most common symptom experienced during treatment was headache. Joint pain was found to decrease during treatment. One person discontinued sofosbuvir plus ribavirin due to myocardial infarction and one discontinued sofosbuvir/velpatasvir due to urticaria.

**Conclusions:** In the real-world setting, sofosbuvir-based treatment is safe, effective, and well tolerated in AN/AI patients. Sustained virologic response was high regardless of HCV genotype or cirrhosis status.

## Introduction

Hepatitis C virus (HCV) infection is one of the main causes of chronic liver disease and liver-related death worldwide (1, 2). The number of chronically infected individuals globally is estimated to be over 130 million (3). Moreover, most new cases of acute HCV infection are not identified or reported to public health authorities because most adults and adolescents with HCV do not have symptoms and are unaware of their infection (4). The U.S. Centers for Disease Control and Prevention (CDC) estimates there were 41,200 new HCV infections in the United States in 2016 alone (4). Chronic hepatitis C is a known risk factor for development of hepatocellular carcinoma (HCC) (5). Out of 700,000 cases of HCC that occurred worldwide in 2008, 17 to 22% are attributable to HCV infection (6, 7).

The prevalence of HCV infection in the United States in 2013-2016 was reported to be 2.4 million cases, with over 30,000 new cases of HCC and approximately 27,000 deaths registered every year (8, 9). The implementation of extended criteria for Hepatitis C screening is a subject of major policy debate underscoring the rising concerns over the growing public health and economic burden of HCV infection (10, 11). The first National Viral Hepatitis Action Plan that aimed to eliminate hepatitis C was issued in 2011(12). It focused on hepatitis B and C elimination by the year 2020 and recognized that success of the action plan could not be achieved without input from non-federal stakeholders, both public and private. Since then, several U.S. states implemented their own hepatitis elimination programs with ambitious goals of eliminating viral hepatitis by 2030, in accordance with the World Health Organization’s directive on eliminating viral hepatitis (13, 14).

A number of studies have described the epidemiology of HCV infection among American Indian or Alaska Native people (15-20). Risk factors for acquisition of HCV infection and genotype (GT) distribution among AN/AI were similar to those found in the National Health and Nutritional Examination Survey (17, 18). End stage liver disease (ESLD) was associated with older age and alcohol usage (20). Previously we found that AN/AI people did not respond well to peginterferon and ribavirin-based treatment with an overall (all GTs) SVR rate of 51% and discontinuation rate of 52% (19). Genotype 3 HCV infection was associated with a high risk for developing ESLD, HCC and liver-related death (18). Bruden et al (20) emphasized the need for early hepatitis C treatment of patients with moderate fibrosis to prevent development of ESLD, HCC and liver-related death over time.

Alaska Native and American Indian (AN/AI) communities are one of the priority populations in the National Viral Hepatitis Elimination Plan (21). A recent study showed that 86% of AN/AI people diagnosed with HCV infection in Alaska were linked to care and treatment in 2017 (16). Despite that, national data show AN/AI people are disproportionately affected by hepatitis C with the highest rates of both acute HCV infection and hepatitis C-related deaths (21).

With the advent of new direct-acting antivirals (DAAs) the HCV treatment landscape has changed. Recent studies report higher efficacy, shorter treatment duration, and fewer side effects with the inclusion of the DAA sofosbuvir into treatment regimens (22-24). Importantly, hepatitis C DAA therapy has eliminated the GT dependent differences in effectiveness that were associated with interferon-based therapies. Public health experts now believe it is possible to eliminate the disease. Hepatitis C elimination could have a tremendous positive effect on improving health outcomes by preventing around 400,000 deaths worldwide from complications each year (25).

There is ongoing discussion about whether the effectiveness of hepatitis C treatment varies between racial and ethnic groups in the United States. The Liver Disease and Hepatitis program (LDHP) at the Alaska Native Tribal Health Consortium (ANTHC) provides a reliable setting to study disparities in hepatitis C treatment due to high rates of the disease in Alaska (26), with an ongoing registry in place, and an established AN/AI HCV-infected cohort from a large and diverse AN/AI population.

In this study we aimed to evaluate the effectiveness of sofosbuvir-based hepatitis C treatment regimens and describe any differences in experience and outcome among AN/AI people living in Alaska as compared to other racial and ethnic groups.

## Materials and Methods

### Study design, setting, participants and data collection

In 1995, the ANTHC LDHP began a hepatitis C population-based cohort study (AK-HEPC) to learn more about adverse outcomes of chronic HCV infection in AN/AI persons. AN/AI persons residing in Alaska can receive care through the Alaska Tribal Health System (ATHS), an integrated system of tribally-owned and operated health care organizations for which ANTHC is the umbrella organization coordinating healthcare throughout the ATHS. The LDHP operates out of the Alaska Native Medical Center (ANMC), the specialty referral hospital for the ATHS located in Anchorage, Alaska. Statewide testing for HCV infection is provided at ANMC for AN/AI people. All persons found to be HCV antibody (anti-HCV) positive are entered into a registry for clinical care. New positive anti-HCV tests are automatically reflexed to HCV RNA testing when specimen quantity permits. Those entered into the registry are invited to join the AK-HEPC cohort study. Sofosbuvir-based treatment for this cohort was studied from March 1, 2014 through September 30, 2019 in 501 patients with chronic HCV.

AN/AI patients within ATHS with confirmed positive anti-HCV and HCV RNA, who were 18 years of age and older, and who provided written informed consent were included in the study. The study was approved by the Alaska Area Institutional Review Board and by two Alaska Tribal Health Organizations, ANTHC and Southcentral Foundation.

Patients were excluded from the study if they had negative HCV RNA prior to treatment, current incarceration, contraindication to taking any component of a sofosbuvir-based regimen, or no indication for treatment in accordance with the American Association for the Study of Liver Diseases (AASLD) and Infectious Disease Society of America (IDSA) HCV Guidance (27).

### Ethical Considerations

Hepatitis C treatment options were discussed with the patient prior to consenting to the treatment study. These options included all available hepatitis C medications, not just sofosbuvir-based drugs. This manuscript examines hepatitis C treatment and outcomes of patients who were treated with sofosbuvir-based drugs only.

AUDIT-C alcohol screening and PHQ-9 depression screening tests were completed by patients prior to treatment start. Those with positive AUDIT-C scores (≥3 for women and ≥ 4 for men) were counseled about hazardous drinking and offered referral to a behavioral health counselor.

### Treatment Regimens

The Food and Drug Administration-approved sofosbuvir-based treatment regimens included in this study were ledipasvir/sofosbuvir, sofosbuvir plus weight-based ribavirin, and sofosbuvir/velpatasvir. Indications and treatment duration in accordance with prescribing information are shown in Table 1.

**Table 1.** Treatment Regimens of 501 AN/AI persons treated for hepatitis C in Alaska using sofosbuvir-based Direct Acting Antivirall therapy, Alaska (2014-2018).

| Treatment Regimen | Duration of treatment<br>and sample size | Indication |
| --- | --- | --- |
| Ledipasvir/sofosbuvir | 8 weeks<br>(n =110) | GT 1 without cirrhosis and HCV viral load < 6 million international units/mL could be given for a shortened course of 8 weeks, in accordance with prescribing information and at the discretion of the prescribing provider |
| Ledipasvir/sofosbuvir | 12 weeks<br>(n =238) | GT 1,4,5,and 6 patients who were treatment naïve without cirrhosis or with compensated cirrhosis or GT 1,4,5, and 6 patients without cirrhosis who had previously been treated with interferon-based treatment |
| Sofosbuvir plus<br>ribavirin | 12 weeks<br>(n = 39) | GT 2 without cirrhosis. Used until sofosbuvir/velpatasvir became available in Aug 2016 |
| Sofosbuvir plus<br>ribavirin | 16 weeks<br>(n = 6) | GT 2 with cirrhosis or previously treated with interferon-based treatment. Used until sofosbuvir/velpatasvir became available in Aug 2016 |
| Sofosbuvir plus<br>ribavirin | 24 weeks<br>(n = 14) | GT 3 regardless of cirrhosis status. Used until sofosbuvir/velpatasvir became available in Aug 2016 |
| Sofosbuvir/velpatasvir | 12 weeks<br>(n = 94) | All GTs without cirrhosis or with compensated cirrhosis starting in Aug 2016 |

Due to low numbers for comparison, those with decompensated cirrhosis treated with sofosbuvir/velpatasvir for 24 weeks (n=1), ledipasvir/sofosbuvir for 24 weeks (n=17), sofosbuvir/velpatasvir with ribavirin for 12 weeks (n=6), ledipasvir/sofosbuvir and ribavirin for 12 weeks (n=9), and ledipasvir/sofosbuvir and ribavirin for 24 weeks (n=4) were not included in this analysis.

Treatment protocol was the same for those who were enrolled in the study cohort and those who chose not to be in the study and based on current recommendations from the AASLD/IDSA HCV Practice Guidance (27). For those participants treated at other mostly rural Alaska Tribal facilities, the same treatment protocol was followed except there was not a requirement to complete a symptoms inventory before and during treatment. For participants treated at other Alaska Tribal facilities, LDHP providers were consulted by rural tribal providers for treatment recommendations.

### Study Assessments

Pretreatment baseline characteristics (Table 2) and laboratory results [including: complete blood count (CBC), comprehensive metabolic panel (CMP), prothrombin time with international normalized ratio, HCV RNA, HCV GT, alpha fetoprotein, vitamin D, glycosylated hemoglobin (HgbA1C), human immunodeficiency virus (HIV), hepatitis B core antibody (anti-HBc), hepatitis B surface antigen (HBsAg), hepatitis A antibody total, IL-28b genotype, and pregnancy test [for women of childbearing age] were assessed. HBV DNA testing was done for those who were found to be anti-HBc positive. Those who were not immune or previously vaccinated for hepatitis A and B were offered the appropriate vaccine series.

**Table 2.** Characteristics of 501 AN/AI persons treated for hepatitis C in Alaska using sofosbuvir-based DAA therapy, Alaska (2014-2018).

| Characteristic | Level | All Persons<br>% (n = 501) | Persons Treated at<br>ANMC<br>% (n = 378) |
| --- | --- | --- | --- |
| Treatment Location | ANMC<br>Non-ANMC | 75% (378)<br>25% (123) | 100% (378) |
| Completed Therapy |  | 96% (482) | 95% (360) |
| Previously treated |  | 5% (27) | 6% (22) |
| IL28b<br>(n= 326) | CC<br>CT<br>TT | 45% (145)<br>46% (151)<br>9% (30) | 45% (120)<br>46% (124)<br>9% (25) |
| Known HIV Positive |  | 2% (8) | 1% (5) |
| Treatment Length | 8 Weeks<br>12 Weeks<br>16 Weeks<br>24 Weeks | 22% (109)<br>74% (372)<br>1% (6)<br>3% (14) | 22% (84)<br>74% (279)<br>1% (4)<br>3% (11) |
| Treatment Regimen | Sofosbuvir+Ribavirin<br>Ledipasvir/Sofosbuvir<br>Sofosbuvir/Velpatasvir | 12% (59)<br>70% (348)<br>18% (94) | 12% (45)<br>67% (252)<br>21% (81) |
| HCV GT | 1a<br>1b<br>2<br>3<br>4, 6 | 65% (327)<br>10% (52)<br>15% (75)<br>9% (45)<br>0.4% (1 each) | 64% (242)<br>10% (39)<br>16% (59)<br>10% (37)<br>0.3% (1 GT 6) |
| Female Gender |  | 46% (229) | 47% (177) |
| PPI/Antacid Use <sup>a</sup> |  | 29% (96/335) | 29% (95/331) |
| Characteristics at Start of Treatment |  |  |  |
| Median Age (range) |  | 54.3 Years (21.3, | 52.3 years (22.3, 72.7) |
| Mean BMI |  | 28.0 | 28.1 |
| % BMI ≥ 30 |  | 29% (122/417) | 30% (106/356) |
| Advanced Fibrosis (F3-F4) |  | 27% (134/497) | 22% (84) |
| Cirrhosis (F4) |  | 14% (69/497) | 13% (49) |
| Median ALT (min, max) at |  | 52 (7, 612) | 51 (7, 612) |
| HCV RNA ≥ 6 million <sup>b</sup> |  | 18% (92) | 20% (76/378) |
| Vitamin D < 30 iu/mL <sup>b</sup> |  | 45% (185/415) | 45% (153/341) |
| Vitamin D < 20 iu/mL <sup>b</sup> |  | 15% (64/415) | 15% (50/341) |
| Mean A1C % (min, max) <sup>b</sup> |  | 5.6 (4.4, 11.6) | 5.4 (4.4, 11.6) |
| Median AFP <sup>b</sup> |  | 3.6 (0.6, 226.1) | 3.4 (0.6, 226.1) |
| Median GFR (mL/min) <sup>b</sup> |  | 94 (38, 257) | 98 (38, 257) |
| Positive Anti-HBc<sup>c</sup>Start |  | 20% (93/460) | 19% (67/360) |

<sup>a</sup> – Defined as use of any PPI, H2 blocker, or magnesium or aluminum containing antacid.
<sup>b</sup> – These labs were collected between 24 weeks prior to treatment start until one week after treatment start. The closest chronologically to the treatment start date was used in calculation of mean or median.
<sup>c</sup> – At any time point prior to treatment

Liver fibrosis was assessed by either vibration controlled transient elastography (VCTE) using FibroScan^®^, serum fibrosis test, or liver biopsy. A pre-treatment VCTE was performed on all patients who were treated through the ANTHC LDHP, except for the first 7 who had begun treatment prior to availability of VCTE at ANTHC. Fibrosis stage was assessed according to Metavir-equivalent scoring (F0 to F4). Median VCTE level ≥12kPa was defined as cirrhosis.

A thorough health history was completed prior to treatment. This included presence or history of the following: any liver disease or cirrhosis; pulmonary disorders; cardiovascular disease; including deep vein thrombosis and pulmonary embolism; use of acid-reducing agents including proton pump inhibitors (PPIs), H2 blockers and antacids; autoimmune disorders; organ transplant; cancer; current infection; high blood pressure; dyslipidemia; kidney disease; anemia; current TB treatment; diabetes Type 1 or 2; HIV; seizure disorder; and mental health conditions. A complete medication history, which included both prescription and over-the-counter supplements, was obtained to identify any potential interactions with sofosbuvir-based drugs. Participants’ vaccine status for influenza, pneumonia, tetanus and Tdap, and shingles were reviewed and those who were not up-to-date on those vaccines were offered them.

The safety and tolerability of antiviral drug regimens were assessed by reviewing the answers on the baseline self-reported symptoms inventory completed prior to treatment and subsequent symptoms inventories which were completed monthly through the end of treatment, to assess for potential adverse effects related to HCV treatment. Symptoms included in the inventory were: feeling excessively tired (fatigue), flu-like illness, chills, fever, weakness, trouble sleeping, headache, hair loss, dry mouth, cough, shortness of breath, decreased appetite, nausea, vomiting, weight loss, heartburn or upset/sour stomach, muscle aches, joint aches, back pain, itching, rash, dizziness, irritability, depression or anxiety, changes in mood/mood swings, and feeling forgetful/problems concentrating.

Prior to treatment start, an HCV treatment agreement was signed by the patient which explained the treatment process, possible side effects, drug interactions, expected efficacy, and asked the patient to agree to the following:

- Not to drink alcohol or use recreational drugs during treatment.
- Report any serious medical conditions, including heart disease, high blood pressure, diabetes, high cholesterol, rheumatoid arthritis, drug addiction, or psychiatric condition.
- Commit to taking medications as prescribed and if unable to do so to contact provider.
- Protect self and others from hepatitis C by not sharing needles, drug works, razors, toothbrushes, or nail clippers, and cover cuts to prevent blood exposure.

Patients were not penalized if they broke the agreement but were encouraged to resume adherence to it. If a patient identified as having a substance use disorder, referral to behavioral health was offered.

Labs obtained monthly during treatment and analyzed included: HCV RNA, CBC, CMP, HBV DNA (if HBsAg positive or if HBV DNA present in those who were anti-HBc positive prior to treatment), and pregnancy test on women of childbearing age.

Treatment completion or discontinuation of medications was documented. Treatment efficacy as determined by sustained virologic response (SVR) 12 weeks after treatment completion was assessed using HCV RNA (COBAS(R) AmpliPrep /COBAS(R) TaqMan(R) HCV Test Kit (Roche Molecular Systems, Inc., Pleasanton, CA). CBC, hepatic function panel and AFP were drawn at SVR time point and VCTE was repeated to assess liver fibrosis at that time.

One year after treatment completion, VCTE was repeated to reassess liver fibrosis. All patients whose pre-treatment fibrosis assessment (either by VCTE, serum fibrosis test, or liver biopsy) indicated advanced liver fibrosis (Metavir F3-F4 equivalent fibrosis) were instructed to continue to have right upper quadrant (RUQ) ultrasound along with AFP blood test every 6 months as surveillance for HCC.

## Data Analysis

For the outcome of DAA treatment success rate, we calculated SVR in an intention-to treat analysis and a per-protocol analysis. The intention-to-treat analysis included all persons who started HCV treatment. The per-protocol analysis removed persons who discontinued treatment early, regardless of their virologic response and persons who were lost to follow-up with an unknown virologic response. We compared the SVR rate using a likelihood ratio χ2 or Cochran-Armitage test of trend. The high SVR rates with an associated low sample size of treatment failures limited the building of multivariable logistic regression models. We used forward selection with log-binomial regression. After inclusion of the most significant univariate variable, no statistically significant multivariable models were found. We compared alanine aminotransferase (ALT), AFP and VCTE results before and after treatment using a normal or log-normal response and generalized estimating equations were used to account for repeated measurements. The prevalence of side effects reported on the symptoms inventory was compared between the start and end of treatment using generalized estimating equations logistic regression. We used the start of treatment as the start of follow-up and the date of HCC detection, death date, or Dec 1, 2019 as the end of follow-up for calculating HCC incidence following treatment. Exact *P*-values were reported when sample size necessitated. All analyses were run in SAS version 9.4 and a *P*-value < 0.05 was considered statistically significant.

## Results

### Characteristics of study participants

Characteristics of patients at baseline are shown in Table 2. The final data analysis occurred on 501 AN/AI persons who received *sofosbuvir*-based treatment from 2014 to 2018. Seventy-five percent of treatments were prescribed by ANTHC LDHP providers on the ANMC campus in Anchorage; (n=378); 25% were prescribed by providers at other Alaska Tribal facilities, following the protocol used at ANTHC LDHP except symptoms inventory was limited to participants treated in Anchorage.

For comparison, study participants were divided into three groups based on the treatment regimen: ledipasvir/sofosbuvir, sofosbuvir plus ribavirin, and sofosbuvir/velpatasvir. Sofosbuvir-based treatment type and duration of treatment were determined by HCV GT, and fibrosis status in accordance with AASLD/IDSA treatment guidance available at the time of treatment (27).

Median age of study participants was 54.3 years, ranging from 21.3 to 78.3. The majority of patients were GT-1a [327 (65%)]; male [272(54%)], and treatment naïve [474(95%)]. The mean body mass index (BMI) was 28.0; 122/417 (29%) patients had a BMI greater than 30. Advanced fibrosis occurred in 134/497 (27%) persons and 69/497 (14%) patients had cirrhosis (F4). HCV RNA greater than six million international units/ml was observed in 92/501 (18%) patients.

Of the 501 persons, 480 (96%) completed their treatment regimen. Twenty percent (93/460) of persons were anti-HBc positive prior to sofosbuvir-based treatment; with 4 of those having chronic hepatitis B (HBsAg positive). Four participants who reported ongoing injection drug use had achieved SVR and then had HCV RNA detected at a later time point, suggesting reinfection. Eight patients (2%) were HIV positive. There were no positive pregnancy tests during the study.

### Therapeutic Response for Sofosbuvir-based Treatment in AN/AI People

Sofosbuvir-based medication consisted of ledipasvir/sofosbuvir (n=-348), sofosbuvir/velpatasvir (n=94), and sofosbuvir plusribavirin (n=59). The most frequent treatment length was 12 weeks (74%), followed by 8 weeks (22%), 24 weeks (3%), and 16 weeks (1%). SVR was analyzed for both intention-to-treat and per-protocol groups in Table 3.

**Table 3.** Treatment success results among AN/AI (sustained virologic response at 12 weeks post treatment) by Intention-to-treat and Per-protocol Analyses with 95% Confidence Intervals [CI], Alaska, 2014-2018

| Category | Level | SVR Intention-to-treat | P-value for Difference | SVR Per-protocol<sup>a</sup> | P-value for Difference |
| --- | --- | --- | --- | --- | --- |
| Overall |  | 95.2% [93.3, 97.1] |  | 97.3% [95.8, 98.7] |  |
| Sex | Female | 96.9% [93.8, 98.8] | 0.09 | 98.2% [95.4, 99.5] | 0.26 |
|  | Male | 93.8% [90.9, 96.6] |  | 96.5% [94.3, 98.8] |  |
| Age | < 40 years | 93.3% [89.7, 98.7] | 0.95 | 99.0% [94.3, 99.9] | 0.18 |
|  | 40-59 years | 95.9% [93.6, 98.3] |  | 97.3% [95.3, 99.3] |  |
|  | ≥ 60 years | 94.5% [90.6, 98.5] |  | 96.0% [92.5, 99.4] |  |
| Birth Cohort | Pre 1966 | 95.0% [92.6, 97.4] | 0.75 | 96.4% [94.3, 98.5] | 0.15 |
|  | After 1966 | 95.6% [92.6, 98.6] |  | 98.8% [95.9, 99.9] |  |
| Treatment Location | ANMC | 95.8% [93.7, 97.8] | 0.32 | 98.3% [96.4, 99.4] | 0.02 |
|  | Rural | 93.5% [89.1, 97.9] |  | 94.2% [88.4, 97.6] |  |
| Completed Treatment | Yes | 97.9% [96.0, 99.1] | <0.0001 | Not Applicable |  |
|  | No | 54.6% [32.2, 75.6] |  | Not Applicable |  |
| Treatment Regimen | Led/Sof<sup>b</sup> | 96.8% [95.0, 98.7] | 0.004<sup>c</sup> | 97.9% [96.4, 99.4]<sup>d</sup> | 0.01<sup>c</sup> |
|  | Sof/Riba | 86.4% [77.7, 95.2] |  | 90.6% [82.7, 98.4] |  |
|  | Sof/Vel | 94.6% [88.0, 98.3] |  | 98.9% [94.0, 100.0] |  |
| BMI ≥ 30 | Yes | 92.6% [88.0, 97.3] | 0.13 | 94.0% [89.7, 98.3] | 0.02 |
|  | No | 96.3% [94.1, 98.4] |  | 98.6% [96.4, 99.6] |  |
| Steatosis (CAP > 248) | Yes | 92.8% [88.7, 96.9] | 0.09 | 94.6% [91.0, 98.2] | 0.05 |
|  | No | 97.0% [94.3, 99.6] |  | 98.7% [99.5, 99.9] |  |
| A1c ≥ 5.7 | Yes | 94.6% [86.7, 98.5] | 0.76 | 97.2% [90.3, 99.7] | 1.00 |
|  | No | 95.5% [93.1, 97.9] |  | 97.8% [96.0, 99.5] |  |
| Hep B Core +<sup>e</sup> | Yes | 93.6% [88.6, 98.5] | 0.40 | 93.3% [88.2, 98.5] | 0.08 |
|  | No | 95.9% [93.9, 97.9] |  | 98.3% [96.9, 99.7] |  |
| Fibrosis Level | F0-F1 | 96.1% [93.9, 98.4] | 0.13 | 99.3% [97.4, 99.9] | 0.002<sup>f</sup> |
|  | F2 | 96.2% [89.2, 99.2] |  | 97.4% [90.8, 99.7] |  |
|  | F3 | 92.3% [83.0, 97.5] |  | 93.4% [84.1, 98.2] |  |
|  | F4 | 92.8% [83.9, 97.6] |  | 92.7% [83.7, 97.6] |  |
| HCV GT | 1a | 95.1% [92.7, 97.4] | 0.14 | 97.4% [95.0, 98.9] | 0.38 |
|  | 1b | 100.0% [93.2, 100.0] |  | 100.0% [93.0, 100.0] |  |
|  | 2 | 94.7% [86.9, 98.5] |  | 95.9% [88.5, 99.1] |  |
|  | 3 | 91.1% [78.8, 97.5] |  | 95.1% [83.5, 99.4] |  |
| HCV RNA Level | ≥ 6 million | 92.4% [87.0, 97.8] | 0.28 | 95.4% [88.5, 98.7] | 0.26 |
|  | < 6 million | 95.8% [93.9, 97.8] |  | 97.7% [96.2, 99.2] |  |
| IL-28b | CC | 97.9% [94.1, 99.6] | 0.22 | 98.6% [95.0, 99.8] | 0.15 |
|  | CT | 94.0% [90.3, 97.8] |  | 95.2% [91.7, 98.7] |  |
|  | TT | 93.3% [77.9, 99.2] |  | 100.0% [87.2, 100.0] |  |
| Compensated Cirrhosis | Yes | 92.8% [83.9, 97.6] | 0.36 | 92.7% [83.7, 97.6] | 0.03 |
|  | No | 95.6% [93.6, 97.5] |  | 98.0% [96.7, 99.4] |  |
| PPI Use <sup>g</sup> | Yes | 92.5% [83.4, 97.5] | 0.35 | 96.8% [89.0, 99.6] | 0.68 |
|  | No | 95.6% [93.7, 97.6] |  | 97.4% [95.8, 98.9] |  |
| Treatment Naive | Yes | 95.6% [93.7, 97.4] | 0.13 | 97.8% [96.4, 99.2] | 0.03 |
|  | No | 88.9% [70.8, 97.7] |  | 88.5% [69.9, 97.6] |  |

<sup>a</sup> – Persons Discontinuing Treatment Were Removed and Persons with Unknown SVR
<sup>b</sup> – *P*-value for Ledsof 8-week (99.1%, 108/109) vs. Ledsof 12-week (95.8%, 229/239) treatment = 0.18
<sup>c</sup> – *P*-value is for Led/Sof and Sof/Vel vs. Sof/Riba
<sup>d</sup> – *P*-value for Ledsof 8-week (99.1%, 108/109) vs. Ledsof 12-week (97.4%, 221/227) treatment = 0.44
<sup>e</sup> – all 4 persons who were positive for HBsAg responded with an SVR at 12 weeks
<sup>f</sup> – *P*-value for F0-F1-F2 vs. F3-F4 :
<sup>g</sup> – 67 persons with omeprazole

First, we looked at SVR in the intention-to-treat analysis. Overall SVR rate for sofosbuvir-based treatment was 95.2% (477/501); for the patients treated at ANMC, SVR was 95.8% versus 93.5% at other Alaska locations. This was not statistically significant. Females did not differ in treatment success compared with males (*P*=0.09). Patients who completed treatment had significantly higher treatment success compared to those who did not finish treatment (*P*<0.0001). Those with cirrhosis responded as well to treatment as those without cirrhosis. Additionally, there were no significant differences in treatment success by age, HCV GT, IL-28b GT, use of acid-reducing agent, anti-HBc positive status, or having undergone a previous HCV treatment.

We next looked at SVR per-protocol (removing those who discontinued treatment and persons lost to follow up following treatment completion). These exclusion criteria reduced the analysis cohort to 479 persons. Overall treatment success improved to 97.3% (466/479). SVR was significantly higher for patients treated at ANMC compared to other Alaska locations (*P*=0.02). Fibrosis level at treatment start became significant to treatment success (p<0.01) in the per-protocol analysis. Age, sex, HCV GT, IL-28b status, or acid-reducing agent use remained non-significant. Higher SVR rate of those without presence of cirrhosis was significant (*P*=0.002) as compared to those with compensated cirrhosis. Also, those who had not undergone previous HCV treatment had a significantly higher SVR rate (*P*=0.03) compared to those who had been previously treated.

Eleven persons developed HCC after their treatment (range of 3 months to 3.7 years after treatment); all had F3 to F4 fibrosis prior to treatment. HCC incidence following DAA treatment was 6.37 cases per 1000 person-years of follow-up.

There were four HCV re-infections (<1%) that occurred in participants who had achieved SVR. All four were found to have a different GT, and had known continued injection drug use. Having resolved hepatitis B infection, isolated anti-HBc positive, or chronic hepatitis B (HBsAg-positive) did not affect SVR rate in our AN/AI cohort. Also, there were no flares of hepatitis B (defined as elevated ALT >2 ULN and HBV DNA > 2,000 IU/mL in persons with previously inactive or resolved HBV) among those who were anti-HBc+ (28), including those with chronic hepatitis B during treatment. There was one death while on HCV treatment due to non-liver related cancer, and two deaths of unknown cause following HCV treatment completion but prior to SVR time point.

#### Changes in Treatment Characteristics from Pre to Post Treatment

We investigated changes in ALT, AFP, and VCTE fibrosis score pre to post treatment for those at ANMC only. All characteristics explored were significant regardless of SVR (Table 4). Treatment success led to reduced AFP levels post treatment and significant reductions in fibrosis scores by VCTE 1-2 years after treatment.

**Table 4.**
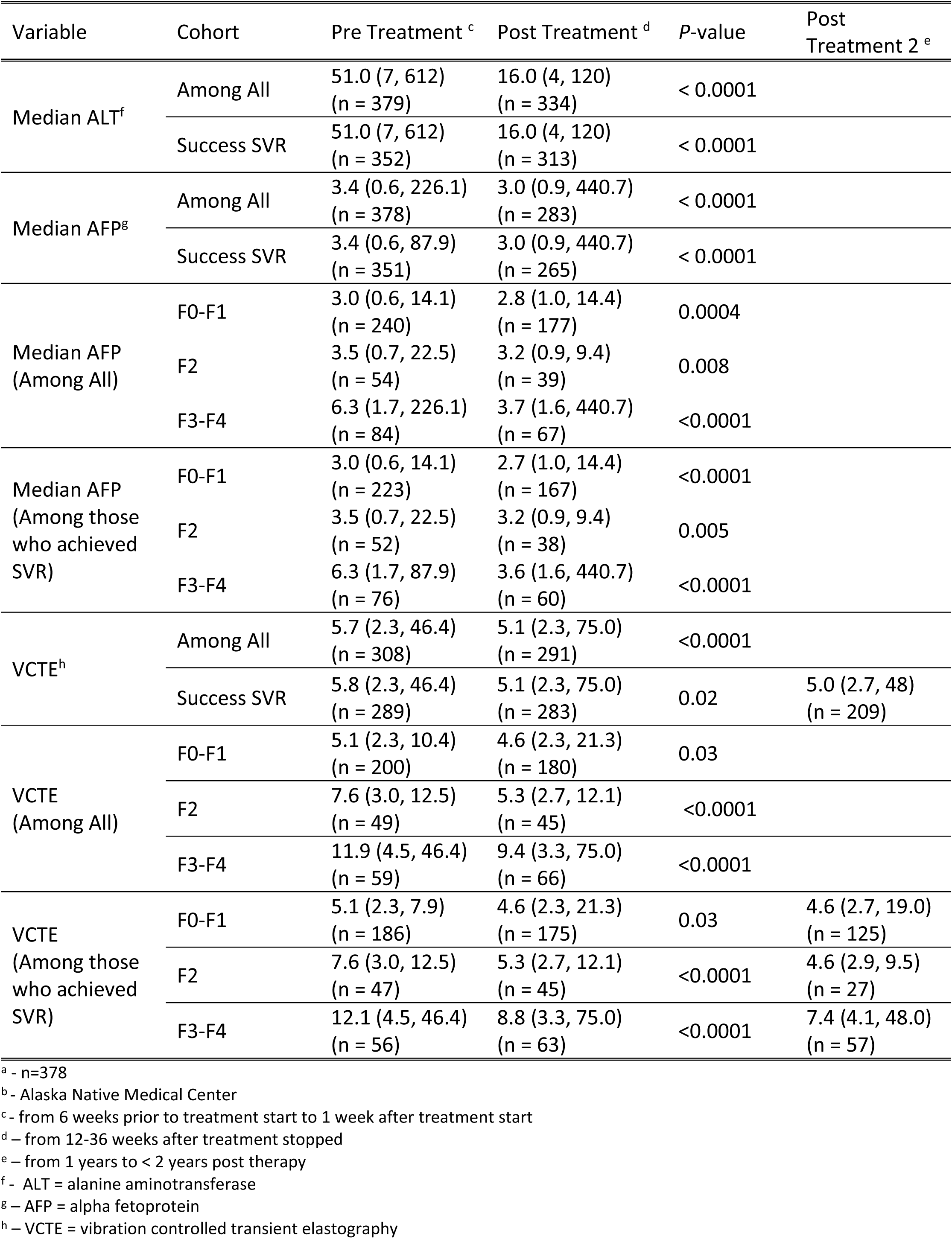
Changes in Fibrosis Markers Among AN/AI Pre and Post Treatment for Persons ^a^ Treated with Sofosbuvir-Based DAAs at ANMC^b,^ Alaska, 2014-2018.

#### Side Effects

Changes in the prevalence of adverse effects, collected through the symptoms inventory, between week 0 and the end of treatment, were analyzed by treatment regimen from the patients treated at ANMC (Table 5). Persons who received sofosbuvir plus ribavirin experienced an increase in mood changes (*P*=0.040), shortness of breath (*P*=0.003), and dizziness (*P*=0.040). Those who received ledipasvir/sofosbuvir reported only an increase in headache (*P*=0.004) on treatment, and those who received sofosbuvir/velpatasvir did not show any statistically significant increases in symptoms during treatment. There was one person who discontinued treatment after one day taking sofosbuvir/velpatasvir because of urticaria. One person discontinued sofosbuvir plus ribavirin after hospitalization for non-ST segment elevation myocardial infarction secondary to coronary vasospasm.

**Table 5.** Symptoms Experienced According to Hepatitis C Treatment Regimen among an Alaska Native/American Indian Cohort^a^ 2014-2018,

| Side Effect<sup>b</sup> | Treatment Week | Drug Regimen |  |  |
| --- | --- | --- | --- | --- |
|  |  | Sofosbuvir/Ribavirin<br>(n= 45) | Ledipasvir/Sofosbuvir<br>(n = 252) | Sofosbuvir/Velpatasvir<br>(n = 81) |
| Tired | 0 | 41% (18/44) | 35% (87/246) | 51% (37/73) |
|  | End of Treatment | 41% (15/37) | 33% (73/221) | 43% (30/69) |
| P-value |  | 0.95 | 0.54 | 0.37 |
| Headache | 0 | 16% (7/44) | 26% (64/246) | 25% (18/73) |
|  | End of Treatment | 22% (8/37) | 36% (79/221) | 25% (17/69) |
| P-value |  | 0.24 | **0.004 ↑** | 0.86 |
| Trouble Sleeping | 0 | 36% (16/44) | 43% (106/246) | 42% (31/73) |
|  | End of Treatment | 49% (18/37) | 35% (78/221) | 38% (26/69) |
| P-value |  | 0.38 | **0.02 ↓** | 0.58 |
| Joint Pain | 0 | 52% (23/44) | 44% (109/246) | 51% (37/73) |
|  | End of Treatment | 27% (10/37) | 32% (70/221) | 38% (26/69) |
| P-value |  | **0.009 ↓** | **0.001 ↓** | **0.04 ↓** |
| Back Pain | 0 | 45% (20/27) | 44% (107/246) | 55% (40/73) |
|  | End of Treatment | 22% (8/37) | 33% (72/221) | 42% (29/69) |
| P-value |  | **0.01 ↓** | **0.002 ↓** | 0.06 |
| Muscle Pain | 0 | 32% (14/44) | 39% (96/246) | 42% (31/73) |
|  | End of Treatment | 24% (9/37) | 31% (69/221) | 33% (23/69) |
| P-value |  | 0.27 | **0.04 ↓** | 0.21 |
| Forgetful | 0 | 23% (10/44) | 31% (76/246) | 32% (23/73) |
|  | End of Treatment | 32% (12/37) | 25% (56/221) | 28% (19/69) |
| P-value |  | 0.30 | 0.12 | 0.64 |
| Dry Mouth | 0 | 18% (8/44) | 24% (59/246) | 15% (11/73) |
|  | End of Treatment | 24% (9/37) | 24% (54/221) | 22% (15/69) |
| P-value |  | 0.39 | 0.76 | 0.14 |
| Weakness | 0 | 9% (4/44) | 18% (45/246) | 30% (22/73) |
|  | End of Treatment | 24% (9/37) | 21% (46/221) | 26% (18/69) |
| *P*-value |  | 0.08 | 0.36 | 0.64 |
| Irritability | 0 | 16% (7/44) | 16% (39/246) | 21% (15/73) |
|  | End of Treatment | 32% (12/37) | 15% (34/221) | 16% (11/69) |
| *P*-value |  | 0.09 | 0.86 | 0.44 |
| Depression | 0 | 11% (5/44) | 25% (62/246) | 34% (25/73) |
|  | End of Treatment | 24% (9/37) | 21% (46/221) | 22% (15/69) |
| *P*-value |  | 0.06 | 0.23 | **0.02 ↓** |
| Itching | 0 | 14% (6/44) | 15% (38/246) | 19% (14/73) |
|  | End of Treatment | 27% (10/37) | 14% (30/221) | 19% (13/69) |
| *P*-value |  | 0.10 | 0.32 | 0.98 |
| Changes in Mood | 0 | 14% (6/44) | 17% (42/246) | 23% (17/73) |
|  | End of Treatment | 30% (11/37) | 17% (38/221) | 16% (11/69) |
| *P*-value |  | **0.04 ↑** | 0.92 | 0.27 |
| Heartburn | 0 | 11% (5/44) | 21% (52/246) | 26% (19/73) |
|  | End of Treatment | 19% (7/37) | 19% (42/221) | 23% (16/69) |
| *P*-value |  | 0.34 | 0.43 | 0.60 |
| Shortness of Breath | 0 | 7% (4/44) | 14% (34/246) | 30% (22/73) |
|  | End of Treatment | 35% (13/37) | 14% (30/221) | 22% (15/69) |
| *P*-value |  | **0.003 ↑** | 0.95 | 0.15 |
| Nausea | 0 | 5% (2/44) | 11% (26/246) | 22% (16/73) |
|  | End of Treatment | 8% (3/37) | 16% (35/221) | 28% (19/69) |
| *P*-value |  | 0.48 | 0.05 | 0.41 |
| Blurred Vision | 0 | 16% (7/44) | 18% (44/246) | 21% (15/73) |
|  | End of Treatment | 19% (7/37) | 15% (34/221) | 15% (10/69) |
| *P*-value |  | 0.71 | 0.42 | 0.36 |
| Decreased Appetite | 0 | 9% (4/44) | 13% (32/246) | 25% (18/73) |
|  | End of Treatment | 14% (5/37) | 13% (29/221) | 9% (6/69) |
| *P*-value |  | 0.53 | 0.97 | **0.009 ↓** |
| Diarrhea | 0 | 5% (2/44) | 8% (19/246) | 14% (10/73) |
|  | End of Treatment | 14% (5/37) | 11% (24/221) | 13% (9/69) |
| *P*-value |  | 0.08 | 0.21 | 0.95 |
| Cough | 0 | 9% (4/44) | 15% (36/246) | 21% (15/73) |
|  | End of Treatment | 16% (6/37) | 12% (27/221) | 20% (14/69) |
| *P*-value |  | 0.32 | 0.43 | 0.95 |
| Chills | 0 | 2% (1/44) | 9% (23/246) | 15% (11/73) |
|  | End of Treatment | 16% (6/37) | 11% (25/221) | 23% (16/69) |
| *P*-value |  | 0.06 | 0.46 | 0.17 |
| Dizziness | 0 | 2% (1/44) | 9% (23/246) | 22% (16/73) |
|  | End of Treatment | 19% (7/37) | 10% (22/221) | 12% (8/69) |
| *P*-value |  | **0.04 ↑** | 0.86 | 0.09 |
| Flu-Like Symptoms | 0 | 9% (4/44) | 7% (16/246) | 11% (8/73) |
|  | End of Treatment | 5% (2/37) | 11% (25/221) | 17% (12/69) |
| *P*-value |  | 0.43 | 0.05 | 0.21 |
| Rash | 0 | 11% (5/44) | 8% (20/246) | 12% (9/73) |
|  | End of Treatment | 15% (6/37) | 7% (16/221) | 10% (7/69) |
| *P*-value |  | 0.44 | 0.65 | 0.83 |
| Hair Loss | 0 | 5% (2/44) | 6% (15/246) | 12% (9/73) |
|  | End of Treatment | 8% (3/37) | 10% (23/221) | 17% (12/69) |
| *P*-value |  | 0.49 | 0.07 | 0.33 |
| Vomiting | 0 | 5% (2/44) | 4% (10/246) | 7% (5/73) |
|  | End of Treatment | 3% (1/37) | 3% (7/221) | 6% (4/69) |
| *P*-value |  | 0.68 | 0.48 | 0.77 |
| Anemia | 0 | 0% (0/44) | 0% (0/246) | 1% (1/73) |
|  | End of Treatment | 3% (1/37) | 3% (6/221) | 0% (0/69) |
| *P*-value |  | Not tested | Not tested | Not tested |
| Fever | 0 | 0% (0/44) | 3% (8/246) | 7% (5/73) |
|  | End of Treatment | 0% (0/37) | 5% (12/221) | 7% (5/69) |
| *P*-value |  | Not tested | 0.24 | 0.93 |

<sup>a</sup> – statistically significant *P*-value change shown in bold with arrow to indicate increase (↑) or decrease (↓) in symptom
<sup>b</sup> - data collected on 33 symptoms

Conversely, after SVR there was a reported significant decrease in joint pain by participants receiving any of the three regimens. Those who received sofosbuvir + ribavirin and ledipasvir/sofosbuvir reported a significant decrease in back pain. Those who received sofosbuvir/velpatasvir reported less depression (*P*=0.02) and less decreased appetite (*P*<0.01). Additionally, those who were treated with ledipasvir/sofosbuvir regimens reported significant decreases in trouble sleeping (*P*= 0.020), joint pain (*P*=0.001), back pain (*P*=0.002), and muscle pain (*P*=0.040).

## Discussion

In this sofosbuvir-based treatment study of AN/AI patients with HCV, overall SVR was achieved by 95.2% of participants. These SVR rates are comparable to rates found in other ethnic groups (29, 30). There were no statistically significant differences in SVR rate by HCV GT, IL-28 GT, or acid blocker use. Sofosbuvir-based HCV treatment regimens were effective and tolerated well by AN/AI persons.

The introduction of all-oral sofosbuvir therapeutic regimens for the treatment of chronic hepatitis C has led to remarkable improvement in SVR, providing therapeutic options to patients with contraindications to IFN-based regimens as well as those who either failed or were intolerant to IFN. However, despite the more convenient dosing schedules and shorter durations of treatment and high cure rates, a small percentage of patients experience treatment failure. The SVR results of our study are in line with earlier studies reporting SVR rates between 87% and 99% (29, 31-34). Furthermore, 92.8% of patients with compensated cirrhosis achieved SVR. The SVR rate for cirrhosis patients achieved in our study is similar to those reported in phase 2 and 3 clinical trials (35, 36).

We assume the high SVR rate of 97.3% in those who completed the study per-protocol was achieved due to high adherence rate and close monitoring and follow-up by experienced nurses and health care providers.

In our study the decreases in non-invasive serological markers (ALT, AFP) and VCTE fibrosis scores suggest a significant regression of inflammation and liver fibrosis in patients at SVR12 as compared to baseline. Several studies have shown the same results; however, the effect was more pronounced in patients with baseline VCTE scores suggesting advanced stages of fibrosis and cirrhosis (37, 38). However, the findings of decreases in liver stiffness scores might be influenced by inflammation, resulting in lower kPa measurements after SVR, which has been previously found in patients with chronic hepatitis (39).

Hepatitis due to reactivation of HBV infection during treatment with DAAs for chronic HCV infection has been reported in patients who were anti-HBc+ prior to treatment. In general, an HBV flare is defined as elevated ALT and HBV DNA > 2,000 IU/mL in persons with previously inactive or resolved HBV infection. Findings from Loggi et al. (40) on 44 subjects who received DAA treatment for HCV infection, found no reactivation in those with past HBV infection (anti-HBc+ only). They also found that two persons who were HBsAg-positive experienced a flare of HBV infection, followed by HBsAg loss. Another study showed that hepatitis B reactivation was a rare event in anti-HBc+/HBsAg-negative patients treated with DAA therapy (41). In our study there were a large number of anti-HBc+ patients (93). Eighty nine of these patients were anti-HBc+/HBsAg-negative whom we assume were infected with hepatitis B and recovered shortly after, and 4 patients were HBsAg-positive, having chronic hepatitis B coinfection. None of these 93 patients experienced flares of HBV during or immediately following treatment. This finding adds support to the body of knowledge showing hepatitis B flares, among those chronically infected and reactivation among those with resolved infection, as a result of HCV treatment, is rare.

HCV reinfection following successful treatment can compromise treatment outcomes. In our study population, we observed less than 1 percent reinfection rate due to ongoing drug use in those who achieved SVR during the span of the study follow up period. A recent meta-analysis showed an overall rate of HCV reinfection of 5.9/100 person-years (95% CI 4.1-8.5) among people with recent drug use (injecting or non-injecting), and 6.2/100 person-years (95% CI 4.3-9.0) among people recently injecting drugs (42).

In our study none of the participants experienced serious adverse effects. The symptoms that increased in frequency were: headache for *ledipasvir/sofosbuvir*, and changes in mood, shortness of breath, and dizziness on *sofosbuvir* plus *ribavirin*, which is likely due to ribavirin. That is in line with previous studies (36, 43-45). Mizokami et al (46) data shows that the most common adverse effects on sofosbuvir-based treatment are nasopharyngitis, anemia, headache and malaise. Difference in the results may be partially explained by the differences in inherent metabolic pathways and different drug-drug interactions.

Our study adds to the body of evidence regarding successful hepatitis C treatment among racial minorities, specifically AN/AI people. It is the largest study to date of American Indian or Alaska Native persons with hepatitis C treated with DAAs (47).

Limitations of our study are that the cohort includes only AN/AI persons, therefore our findings may not apply to other racial/ethnic groups. However, this cohort was described previously and found to closely represent the US population infected with HCV in terms of HCV GT distribution and risk factors for infection (17).

Treatment of HCV infection does not only benefit those infected but also reduces the burden of hepatitis C at the population level (2). Regardless of stage of liver disease, coinfection, or complications there is a survival benefit of SVR (2). Cure of HCV infection has a long term impact on the economic burden of disease (48). Complications of HCV infection such as liver cirrhosis, HCC and transplantation are associated with high medical cost and use of healthcare resources (49). From a public health perspective, the “treatment as prevention” model as undertaken by ANTHC LDHP is of great benefit. It may reduce the prevalence of HCV infection in the population in the long-term, leading to a lower transmission rate and incidence of disease. To achieve 2030 targets of a 90% reduction in new cases of HCV infection and a 65% reduction in liver-related death, a new capacity for screening and treatment should be built. Hepatitis C needs to become a disease of high priority in both clinical and public health settings.

## Conclusion

Our treatment study with sofosbuvir-based regimens was effective for a broad range of AN/AI patients with HCV GT 1, 2, 3, 4, or 6 infection. The treatment was also effective among patients with cirrhosis. These real-life study findings, in a population not included in pre-licensure clinical trials, suggest that HCV treatment for AN/AI persons should lead to a long-term decrease in liver disease progression and prevention of the development of HCC and reduction in disparities of HCV liver disease for AN/AI people.

To date, over 1000 AN/AI persons in Alaska have been treated through the ATHS and cured of hepatitis C with DAA therapy. However, there remains another 1000 known AN/AI persons in Alaska with hepatitis C including approximately 180 AN/AI persons living in over 100 isolated communities without primary care providers and not connected to the road system. Moving forward, the use of telemedicine and other innovative ways to test and treat HCV are being explored so that all AN/AI people can access antiviral treatment, disparities in liver disease outcomes can lessen, and the overarching goal of HCV elimination can be met in the AN/AI population.

## Data Availability

Access to data for this study is subject to tribal review. Requests for data should be directed to the Alaska Native Tribal Health Consortium Privacy Officer c/o Ethics and Compliance Services, 4315 Diplomacy Drive, Anchorage, AK 99508.

## Disclaimer

Dana Bruden is employed by the CDC. The findings and conclusions in this study are those of the authors and do not necessarily represent the official position of the CDC.

